# Using machine learning to evaluate the value of genetic liabilities in classification of hypertension within the UK Biobank

**DOI:** 10.1101/2024.03.18.24304461

**Authors:** Gideon MacCarthy, Raha Pazoki

## Abstract

**Background and objective:** Hypertension increases the risk of cardiovascular diseases (CVD) such as stroke, heart attack, heart failure, and kidney disease, contributing to global disease burden and premature mortality. Previous studies have utilized statistical and machine learning techniques to develop hypertension prediction models. Only a few have included genetic liabilities and evaluated their predictive values. This study aimed to develop an effective hypertension prediction model and investigate the potential influence of genetic liability for risk factors linked to CVD on hypertension risk using Random Forest (RF) and Neural Network (NN).

**Materials and methods:** The study included 244,718 participants of European ancestry. Genetic liabilities were constructed using previously identified genetic variants associated with various cardiovascular risk factors through genome-wide association studies (GWAS). The sample was randomly split into training and testing sets at a 70:30 ratio. We used RF and NN techniques to develop prediction models in the training set with or without feature selection. We evaluated the models’ discrimination performance using the area under the curve (AUC), calibration, and net reclassification improvement in the testing set.

**Results:** The models without genetic liabilities achieved AUCs of 0.70 and 0.72 using RF and NN methods, respectively. Adding genetic liabilities resulted in a modest improvement in the AUC for RF but not for NN. The best prediction model was achieved using RF (AUC =0.71, Spiegelhalter z score= 0.10, P-value= 0.92, calibration slope=0.99) constructed in stage two.

**Conclusion:** Incorporating genetic factors in the model may provide a modest incremental value for hypertension prediction beyond baseline characteristics. Our study highlighted the importance of genetic liabilities for both total cholesterol and LDL within the same prediction model adds value to the classification of hypertension.

## Introduction

Approximately, 1.28 billion people aged 30 to 79 have hypertension worldwide (1) and it continues to rise globally causing a significant socioeconomic burden due to low awareness and poor control (2). Hypertension significantly increases the risk of cardiovascular diseases including stroke, heart attack, heart failure, and kidney disease, contributing to the global disease burden and premature mortality (1, 3, 4).

Every year, the burden of hypertension and related cardiovascular diseases is increasing in the United Kingdom (UK). As of 2017, hypertension prevalence in England was estimated at around 26.2% among adults (5). It is responsible for more than half of all strokes and heart attacks, costing the National Health Service (NHS) more than £ 2.1 billion per year (6).

The current guidelines (7–9) suggest lifestyle modification and the use of blood pressure-lowering medication for preventing hypertension and its consequences. Medication is often successful in lowering blood pressure and reducing the risk of hypertension-related cardiovascular disease and stroke. Lifestyle modifications also offer benefits like reduced drug costs, improved control of other disorders like diabetes and hypercholesterolemia, and avoiding unwanted pharmacological therapy (10). The current guidelines have remained silent on the genetic components of hypertension, which are quantifiable at birth and may be used to determine an individual’s lifelong disease risk before clinical risk factors are established (11), allowing adequate time to determine lifetime measures to lower hypertension risk, particularly in a high-risk group.

Genome-wide association studies (GWAS) have identified numerous multiple single nucleotide polymorphisms (SNPs) associated with hypertension and/or high blood pressure levels (12–17). Developing methods to incorporate genetic factors into prediction models of hypertension has the potential to improve hypertension prediction, management and control.

Previous studies have successfully predicted hypertension using standard statistical techniques or machine learning (18–25). A study in rural Chinese populations (25) incorporated a single hypertension polygenic risk score and showed improvement in hypertension prediction using several machine learning techniques. Several studies (26–28) have used a method called metaGRS to combine several polygenic risk scores into regression models for cardiovascular disease and showed that including multiple genetic factors improves the prediction model’s accuracy compared to using one genetic liability. Whether machine learning methods could be used to improve prediction models from multiple genetic liabilities remains to be elucidated.

Recent studies have provided evidence for genetic correlations between hypertension and type 2 diabetes (29), adiposity traits, (30), lipids traits (31, 32) and smoking traits (33). In the current study, we created genetic liabilities using these risk factors and used machine learning models to test the best combination of genetic liabilities and clinical factors that could optimise the prediction of hypertension in the European ancestry population.

## Material and method

### Ethical Approval

We received approval for this study from the UK Biobank Research Ethics Committee and Human Tissue Authority, and all the participants gave informed consent. This study is done using the UK Biobank data under application number 60549. Additionally, we obtained ethics approval from Brunel University London, College of Medicine, and Life Sciences Research Ethics Committee to work with secondary data from the UKB (reference 27684-LR-Jan/2021-29901-1).

### Study Population

The UK Biobank (UKB) is a prospective observational study with more than half a million participants aged between 40 and 69 years. The participants were recruited between 2006 and 2010 across 22 centres located throughout the United Kingdom (UK). The full description of the UKB study as well as the data collected and a summary of the characteristics, are publicly available on the UKB website (www.biobank.ac.uk) and elsewhere by Sudlow and colleagues (34). In brief, during the recruitment, detailed information about socio-demographics, health status, physician-diagnosed medical conditions, family history, and lifestyle factors was collected via questionnaires and interviews. Several physical measurements, including height, weight, body mass index (BMI), waist-hip ratio (WHR), systolic blood pressure (SBP), and diastolic blood pressure (DBP) were obtained. The records of participants in the UKB project were accordingly linked to Hospital Episode Statistics (HES) data, as well as national death and cancer registries.

The current study is based on a subset of unrelated individuals of European ancestry (n=244,718; Figure 1). In brief, we used 40 genetic principal components created centrally by the UKB and applied the k-means clustering method on 502,219 UKB participants to identify individuals of European descent. We then obtained genetic data from the individuals who had passed the UKB internal quality control and had genotype data (n=459,042). We excluded participants who had withdrawn their consent (n=61), mismatched genetic and self-reported sex (n= 320), were pregnant, or were not sure of being pregnant(n=278). Using the kinship cut of 0.0884 for third-degree relatives, we excluded participants who were up to second-degree related (n=33,369). We further excluded participants who were diagnosed with a stroke, heart attack, or angina before or at baseline (n=25,340), and participants with missing data on the potential confounders (n=61,961; see statistical analysis for detail). Furthermore, individuals who were on cholesterol-lowering medication (n=34,243), stopped smoking or drinking due to health reasons, or doctor’s advice (n=58,752) were excluded from the dataset leaving a final 244,718 unrelated individuals of European ancestry for our analyses.

**Figure 1.**
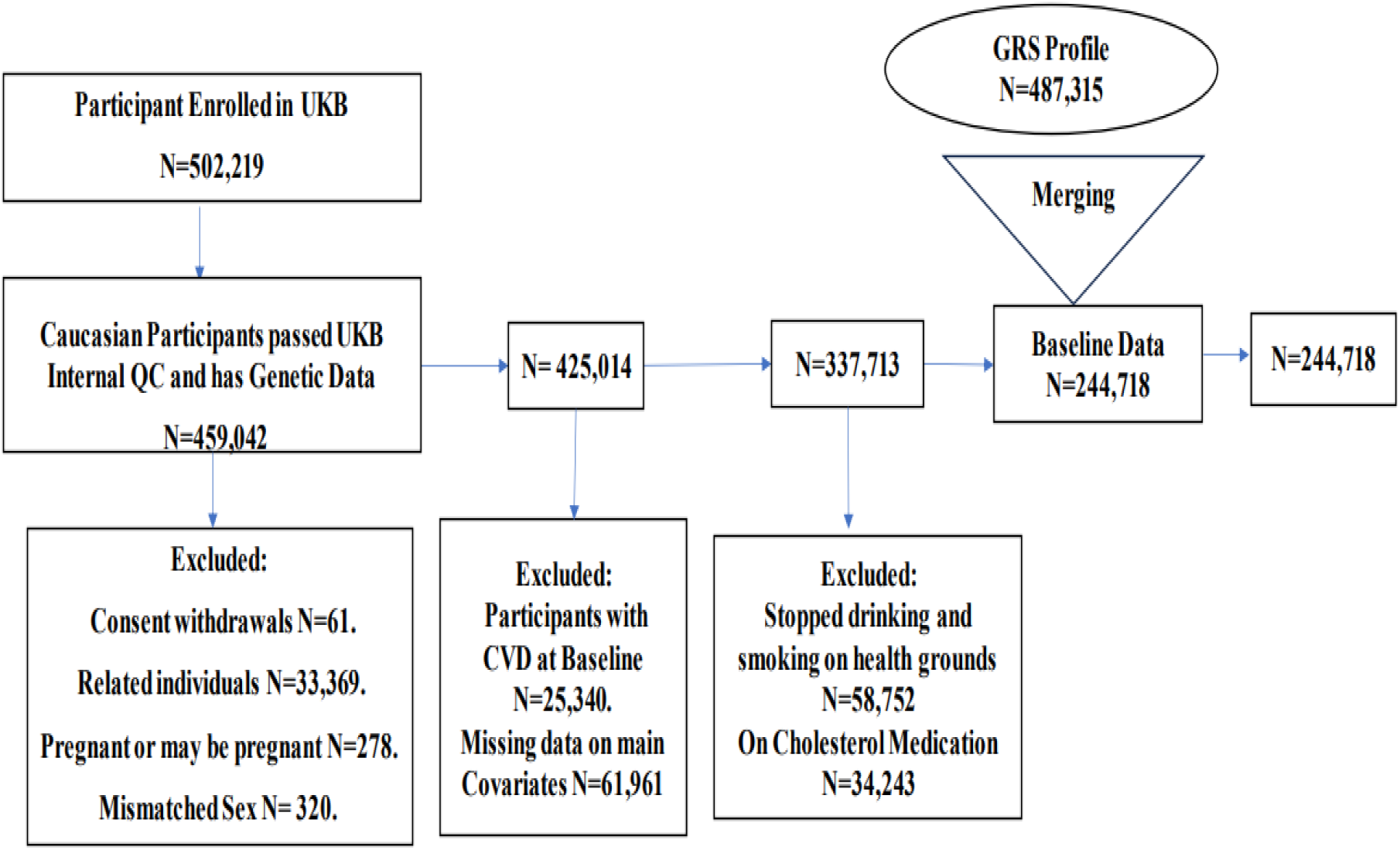
Exclusion Criteria of the study: The flowchart of the study participant selection. UK biobank (UKB) data had 502,219 participants at the time of beginning of this study. We extracted 459,042 participants of European ancestry who have passed UKB internal quality control (QC) and have genetic data. The final dataset included 244,718 participants who have met the inclusion criteria.

### Genotyping and imputation

The UKB conducted all the DNA extraction, genotyping, and imputation. The detailed processes have been discussed elsewhere (35–37). In brief, blood samples from participants were obtained at UKB assessment centers, and DNA was extracted and genotyped using the UKB Axiom Array. The genotype imputation was conducted by UKB using the IMPUTE4 tool. Three reference panels: Haplotype Reference Consortium, UK10K, and 1000 Genomes phase 3 were used for the imputation. The genetic principal components and kinship coefficients were calculated centrally by UKB to account for population stratification and identify related individuals (35, 37).

### Variables and outcome

#### Definition of the outcome

Our main outcome is hypertension which was defined as (1) the presence of a recorded SBP ≥ 140 mmHg or a DBP ≥ 90 or (2) hypertension diagnosed by a doctor or (3) a record of using blood pressure (BP) lowering medication at baseline (38). In the UKB, two blood pressure readings were obtained a few minutes apart using a standard automated device or manual sphygmometer (www.ukbiobank.ac.uk). We calculated both mean SBP and mean DBP from two automated or two manual readings of BP measurements. For Participants with one manual and one automated BP reading, the average of these two values was used. For individuals with a single BP measurement (one manual or one automated BP reading), the single measurement was used for approximating the participant’s BP value. For the participants who self-reported to be taking BP-lowering medication, we added 15 mmHg to SBP and 10 mmHg to DBP (39). The participants with missing BP readings were excluded.

#### Demographics, Clinical and lifestyle features

We included factors such as age, sex, BMI, diabetes mellitus, total cholesterol (TC), Low-Density Lipoprotein (LDL), High-Density Lipoprotein (HDL), smoking status, drinking status, and sedentary lifestyle in our study. Diabetes was defined as a record of diabetes diagnosed by a doctor or using insulin medication or a record of serum level of hemoglobin A1c (HbA1c) ≥ 48 mmol/mol (6.5%) or glucose level ≥ 7.0 mmol/dl (40). Smoking and alcohol consumption data were collected through a self-reported questionnaire by the UKB and were classified into current, previous, and never.

We calculated a sedentary lifestyle variable by approximating the total self-reported hours per day the participants spent on (1) driving, (2) using a computer, and (3) watching television. We considered 30 minutes of sedentary behavior if individuals indicated that they spent less than an hour per day driving or watching television or using a computer.

### Computation of genetic liabilities

#### Single Nucleotide Polymorphism (SNP) Selection

We selected a list of genetic variants in the form of SNPs (table 1) that were previously identified as associated with 10 cardiovascular risk factors traits including three smoking traits, four lipid traits, two adiposity traits, and type 2 diabetes at a genome-wide association study (GWAS) significant threshold (*P*-value <5.0×10^−8^) in the European population. We included SNPs from studies mentioned in Table 1 ensuring no overlap with the UKB to avoid bias. SNPs that were palindromic or in high linkage disequilibrium (LD; r^2^ <0.1) were removed by LD pruning which was performed using the SNPclip module of the LDlink (https://ldlink.nci.nih.gov; access date: 15 /05/ 21). In brief, LDlink is a suite of web-based applications, an LD analysis tool, which is designed specifically for easy and efficient examination of linkage disequilibrium between a set of SNPs in population groups (41). The SNPs used in calculating the GRSs were pruned with the LD pruning procedure employed in LDlink using a threshold of minor allele frequency (MAF) = 0.01 and r^2^ = 0.1. The SNPclip module removed SNPs if they were duplicates, not biallelic, had MAF < 0.01, and were in LD with other SNPs (r^2^ >0.1).

**Table 1:** Characteristics of the genetic variants included to create each genetic liability variable within the UK biobank.

| Trait category | Genetic liability | Study (publication year) | Number of SNPs | reference |
| --- | --- | --- | --- | --- |
| Smoking | Smoking initiation | Liu et al. 2019 | 311 | Liu et al., 2019 |
|  | Smoking cessation | Liu et al. 2019 | 16 | Liu et al., 2019 |
|  | Smoking Heaviness | Liu et al. 2019 | 38 | Liu et al., 2019 |
| Diabetes | Type 2 diabetes | Mahajan et al. 2018 | 210 | Mahajan et al., 2018 |
| Adiposity | BMI | Winkler et al. 2016 | 159 | Winkler et al., 2016 |
|  | WHR | Shungin et al., 2015 | 39 | Shungin et al., 2015 |
| Lipid traits | Total cholesterol | Surakka et al., 2015 | 36 | Surakka et al., 2015 |
|  | HDL | Surakka et al., 2015 | 19 | Surakka et al., 2015 |
|  | LDL | Surakka et al., 2015 | 30 | Surakka et al., 2015 |
|  | Triglycerides | Surakka et al., 2015 | 25 | Surakka et al., 2015 |

SNPs: Single nucleotide polymorphisms, BMI: Body mass index, WHR: waist-hip-ratio, HDL: High density lipoprotein, LDL: Low density lipoprotein

We used the final list of selected LD-pruned SNPs to estimate genetic liabilities for all the 10 traits under the current study using PLINK version 1.9 (42). To allocate weight to each SNP, we used the effect sizes estimated for the association of the SNPs with each of the traits mentioned in Table 1. These effect estimates were obtained from previously published, publicly available genome-wide association summary statistics data (Supplementary Data 1-10). PLINK uses a weighted method where the effect size (beta coefficient) of each SNP is considered as weight and is multiplied by the number of risk alleles an individual carries. The product is then summed across all SNPs to produce genetic liability for each person. All the genetic liabilities were standardized (i.e. mean-centered with standard deviation 1).

### Statistical Analysis

We summarized the categorical variables using frequencies and percentages and the continuous variables were expressed as mean (SD) in Table 3. When comparing the characteristics differences between hypertension and non-hypertensive groups, the nonparametric test (Wilcoxon rank sum test) was utilized for continuous variables. The Chi-square test was used to compare hypertension and non-hypertensive groups for categorical variables. We also performed univariable and multivariable logistic regression analysis to assess the relation between the outcome and the selected features and present the results in supplementary tables1and 2. The statistical significance was defined where associations demonstrated a 2-sided P-value less than 0.05 (Supplementary Tables1 and 2).

### Training and testing datasets

In this study we employed the train-test split approach (43), we randomly partitioned the dataset at a ratio of 70:30 (Figure 2) into a training set (70%; n=171,304; case =81,967 and control = 89,337) and a testing set (30%; n=73,414) using the “*createDataPartition*” function in the R-package. The training set is used to train the models, whilst the testing set is reserved for the final model evaluation. To ensure that all the numerical variables contribute equally to our models (44), we applied the max-min scaling method to the numerical variables to bring them to a common scale before training the models. This is because some of the numerical variables were measured on different scales.

**Figure 2.**
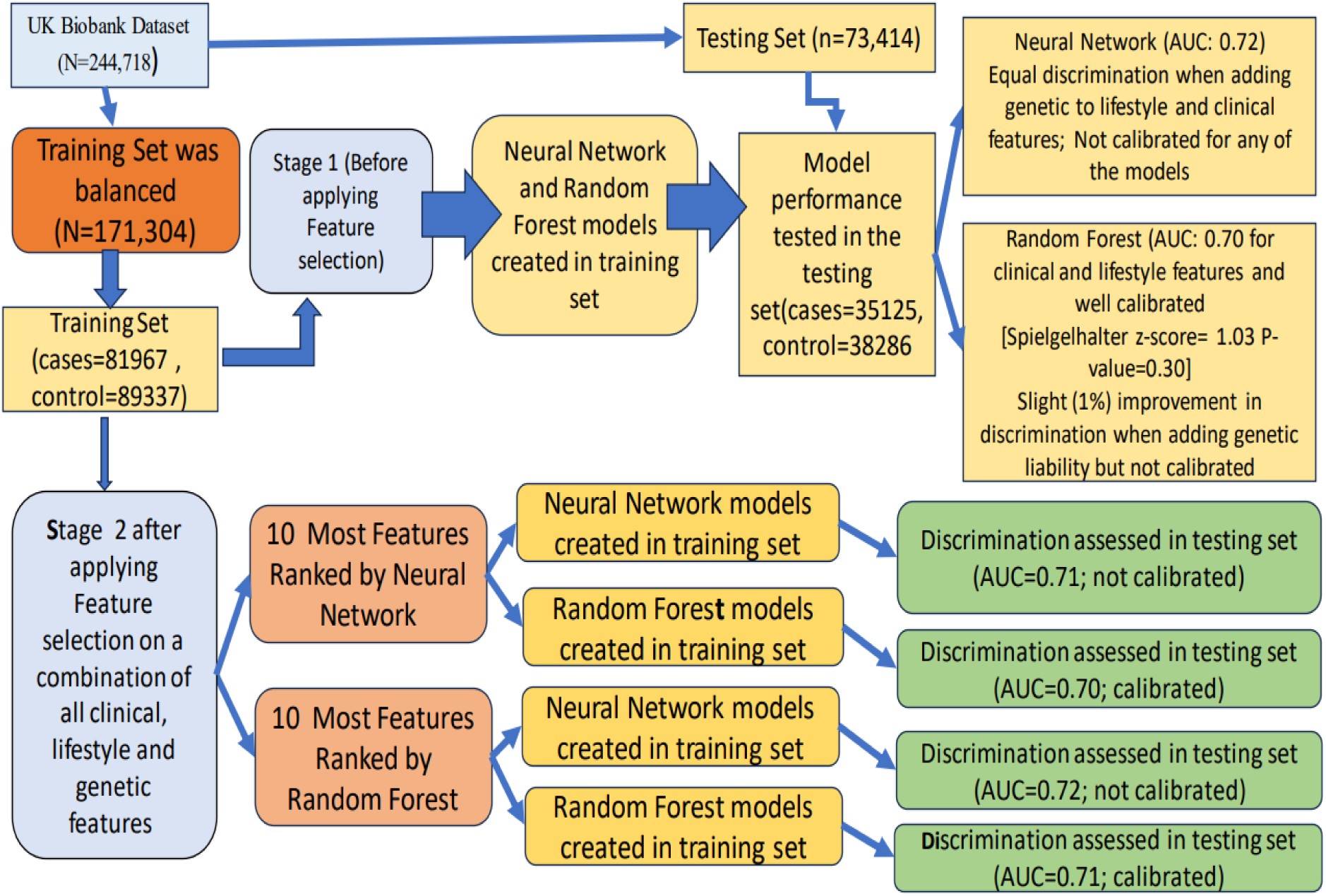
Overview of the study and the construction of machine learning models: The flowchart of the study design. The data was split into training and testing sets. Stage 1 models were constructed in the training set without selection and their performances assessed in the testing set. Stage two models were built after feature selection was applied and their performances evaluated in the testing set.

The models were trained in the training set and the performance of the models in terms of discrimination ability (defined as the model’s capacity to distinguish between persons with and without outcomes) was assessed in the testing set (n=73,414; Figure 2). To this end, we constructed the receiver operating characteristic curve (ROC) for each model and calculated the area under the curve (AUC) with 95% confidence intervals (Cl) (45–47). The AUC ranges from 0.5 to 1.0, with 0.5 indicating no better discrimination than chance and 1.0 representing perfect discrimination power.

### Handling Data Imbalance

Models trained on imbalanced datasets may become biased towards the dominant class, predicting the minority class incorrectly (48). A binary classifier, which is the case in the current study, trained on a balanced dataset typically outperforms a model trained on an imbalanced dataset (49). The imbalanced class in the training set can lead to an unjustified evaluation of two-class classification algorithms. To overcome this issue, we utilized the random over-sampling method using a bootstrapping method to balance the number of events in the training set before training the models. To do this the “*ROSE”* (Random Over-Sampling Examples) package (50) included in the R-program was utilized. The package deals with imbalanced datasets by generating synthetic samples for the minority class to balance the class distribution (50).

### Machine learning model construction

Machine learning algorithms, unlike traditional statistical techniques, are flexible and free of prior assumptions, such as the type of error distribution, capable of capturing the complicated, nonlinear relationships between predictors. It is an application of algorithms to automate decision-making processes using models that have been trained on historical data (51).They can analyze various data types and integrate them into predictions for disease risk (52). There are many types of machine learning algorithms, but some of the most common ones include Support vector machines (SVM), Decision tree (DT), Random Forest (RF), and Neural Network (NN). Prior studies have investigated SVM, RF DT and XGBoost in prediction of hypertension in European samples (53). In this study, we considered two machine learning-based classifiers, the Random Forest, and Neural Network, which have been shown in many studies to have been most promising in classification of hypertension among individuals of European ancestry (53).

The Random Forest is a powerful machine learning algorithm that constructs an ensemble or forest of decision trees that are often trained using the bagging method. Each decision tree is constricted on a random subset of the training set and a random subset of the features. This keeps the trees from getting overly correlated and hence overfitting the data (54). In this study, the Random Forest models were constructed with the “*ranger*” package in the R-program (55) without hyperparameter tuning (all the hyperparameters were set to the default setting on the “*ranger”* package i.e. 500 trees and 10 nodes) and the optimal model was selected based on out-of-bag (OOB) estimate of the error rates in the training set. In a Random Forest model, the maximum number of features that can be considered for splitting at each node of the decision trees within the ensemble was determined and reported using the “*mtry*” parameter within the “*ranger”* package (Supplementary Table 3).

Neural Network is another powerful machine learning algorithm that automatically learns from patterns between the inputs and the output within the data (56). The Neural Network consists of interconnected processing nodes organised in three layers: input, hidden, and output layers (Supplementary Figure S4). The input layer is connected to the hidden layer with updated weight, which is then connected to the output layer (57).

Overfitting and underfitting are two common problems in machine learning that can have a major impact on the performance and generalization ability of models (58). Overfitting is where the model fits the training set properly but performs poorly on the testing or unseen dataset thereby resulting in low training error, but high-test error. Underfitting is simply the opposite of overfitting.

To prevent the problem of overfitting, and underfitting within the Neural Network models, and to provide more reliable estimates of the predictive ability of the models on unseen data (59, 60), we performed a 5-fold cross-validation on the balanced training set(n=171,304) and the receiver operation characteristic (ROC) (61) was used to select the optimal model (the largest ROC value). The optimal threshold for the ROC value from 5-fold cross-validation was identified as 0.70 (Supplementary Table 4). This means that models with ROC values above 0.70 in the testing set would improve prediction. We constructed the Neural Network classifiers with the “*nnet*” function (62) implemented in the R-program “*caret*” package.

In the current study, we adopted a two-stage approach in the construction of the machine learning algorithm.

#### Stage one models

In stage one, we built models without feature selection in the training set (n=171,304). In the first subset of models, we used Random Forest method (55, 63) and in the second subset of models, we used Neural Network (**Figure 2**). For each of these methods, we used two different sets of features. (1) the traditional features that included baseline characteristics (i.e. age, sex, BMI, diabetes mellitus, smoking status, drinking status, total cholesterol (TC), HDL, LDL, and sedentary lifestyle). (2) Saturated features included all baseline characteristics above together with additional genetic variables including ten genetic liabilities.

The optimal number of features used for splitting at each of the decision trees was identified as 3 features in the construction of traditional Random Forest model and this was identified as 4 features in the construction of the saturated Random Forest model. Both Random Forest models above showed a prediction error of 0.22 (Supplementary Table 3).

In the construction of traditional and saturated Neural Network models, the optimal number of hidden layers was identified as 5 hidden layers.

In the testing set, we tested and compared the performance of (1) the Random Forest model with genetic liabilities and the Random Forest model without genetic liabilities (saturated vs. traditional model). (2) the Neural Network model with genetic liabilities and the Neural Network model without genetic liabilities (saturated vs. traditional model).

#### Stage two models

In stage two, we built models with a feature selection step. Similar to stage one, we used Random Forest and Neural Networks as our classification method. We applied the feature selection method on the variables to achieve the best model performance and to minimize capturing unnecessary noise or random fluctuation in the data and to overcome the problem of overfitting that may be caused by irrelevant features. Feature selection was achieved using the Random Forest and Neural Network methods and selected the top ten most important features from the list of all the twenty features under the study. The most important features were extracted with “*vip*” functions in the R-program. Features were ranked based on their predictive power (figure 1). We then used the topmost important features to build models using both Random Forest and Neural Network as the classifying method regardless of the method used in feature selection step. This approach created four different analysis paths to hypertension classification including path (1) where the feature selection model was Random Forest, and the classifying method was Random Forest as well; path (2) where Random Forest was used as the feature selection method and classification method was Neural Network; path (3) where feature selection model was Neural Network, and the classification method was Neural Network as well; path (4) where Neural Network was feature selection method and classification method was Random Forest (see Figure 2). In the testing set, we used the AUC (see above) to assess the performance of these four models built with the 10 most important features selected. Stage one and stage two resulted in construction and testing of a total of eight models.

### Model Performance Assessment by Calibration

We used a calibration curve and Spiegelhalter z score test to examine the models’ calibration (64, 65). Model calibration measures the ability of a model to accurately predict an outcome (66, 67). In the calibration curve, the Y-axis represents the observed probability, and the X-axis represents the predicted probability of developing a disease. The calibration curve includes a diagonal line (i.e. Ideal line) that indicates the prediction of the ideal model. A model is said to be well calibrated if the calibration curve stays close to the line of perfect calibration (i.e. a 45-degree line with an intercept of 0 and a slope of 1). Overestimation and underestimation are represented by curves below and above the ideal calibration line, respectively. The Spiegelhalter z test is a statistical test used to assess the calibration accuracy of a risk prediction model. A perfectly calibrated model would have a Spiegelhalter z score of zero. A Spiegelhalter z score close to zero indicates good calibration, while a Spiegelhalter z score far from zero indicates poor calibration. A positive Spiegelhalter z score indicates that the model is over-calibrated, meaning that the predicted probability of the outcome is too high. A negative Spiegelhalter z score indicates that the model is under-calibrated, meaning that the predicted probability of the outcome is too low.

To confirm the overall accuracy of the models, we also calculated the Brier score (68), which is the mean square error (MSE) between observed and predicted outcomes. The Brier score evaluates both the calibration and discrimination ability of a model (67). The scores range from 0 to 1, with lower scores suggesting superior calibration. Brier scores approaching 0 imply that the model has been adequately calibrated and discriminated. We used the *“val. prob”* function from the *“rms”* packages in the R-program to generate calibration curves, Spiegelhalter z test, and Brier score.

#### Net reclassification index and Integrated discrimination index

We conclusively assessed the performance of well calibrated models using the net reclassification index and integrated discrimination index statistics. The net reclassification improvement is a commonly used metric to compare the relative ability of two models to classify individuals as low- and high-risk (69). A positive net reclassification index value indicates that the new model correctly reclassifies more individuals into higher or lower risk categories compared to the old model. Conversely, a negative net reclassification index value suggests that the old model is better at reclassifying individuals than the new model.

The integrated discrimination index statistic is used to measure the improvement in the ability of two models to distinguish between event and nonevent (70, 71). A positive integrated discrimination index value implies an improvement in the model’s discriminative ability, while a negative integrated discrimination index value suggests a deterioration in the discriminative ability of the new model. In this study, we used the *“reclassification”* function from the *“PredictABEL”* packages in the R-program to obtain the integrated discrimination index and integrated discrimination index values. The discrimination ability, calibration, and reclassification results are described further in Figure 2, Tables 4 and 5. All the analysis was performed with (R version 4.2.2, www.r-project.org), and for reproducibility, we set the seed of random number generator to a value 500 throughout this analysis.

## Results

### Baseline Characteristics of the Participants

A total of 244,718 (Figure 1) unrelated individuals of European ancestry from the UK Biobank were included in this study (Table 3). The average age of the participants was 55.4 ±7.98 years old and 141,931 (58.0%) were female. The sample contained 7011 (2.9%) participants with diabetes. The majority (n= 229,539(93.8%) of the participants reported to be current alcohol drinkers and 164,847 (67.4%) reported to have never smoked. The average BMI was 26.8 (4.58) kg/m2. The sample included 117,095 (47.8%) participants with hypertension. More women were hypertensive then men (52.4% vs. 47.6%; P-value<0.001). The hypertensive participants were older (57.6 ± 7.53 vs. 53.4 ± 7.84 years; P-value<0.001), had higher BMI (28.0 ± 4.83 kg/m^2^ vs. 25.8 ± 4.06 kg/m^2^; P-value<0.001), had higher total cholesterol levels (6.05 ±1.06 vs. 5.79 ± 1.04 mmol/L; P-value<0.001), and had more sedentary lifestyle (4.87 ± 2.39 vs. 4.50 ± 2.33 hours per day; P-value<0.001) than the non-hypertensive participants. There were statistically significant differences in all baseline characteristics between the hypertensive and non-hypertensive groups (Table 3). A total of 20 clinical and genetic features were included in the analysis (see methods; Table 2). All the demographic, clinical and lifestyle features had a statistically significant association with hypertension (Supplementary Tables 1 and 2).

**Table 2:**
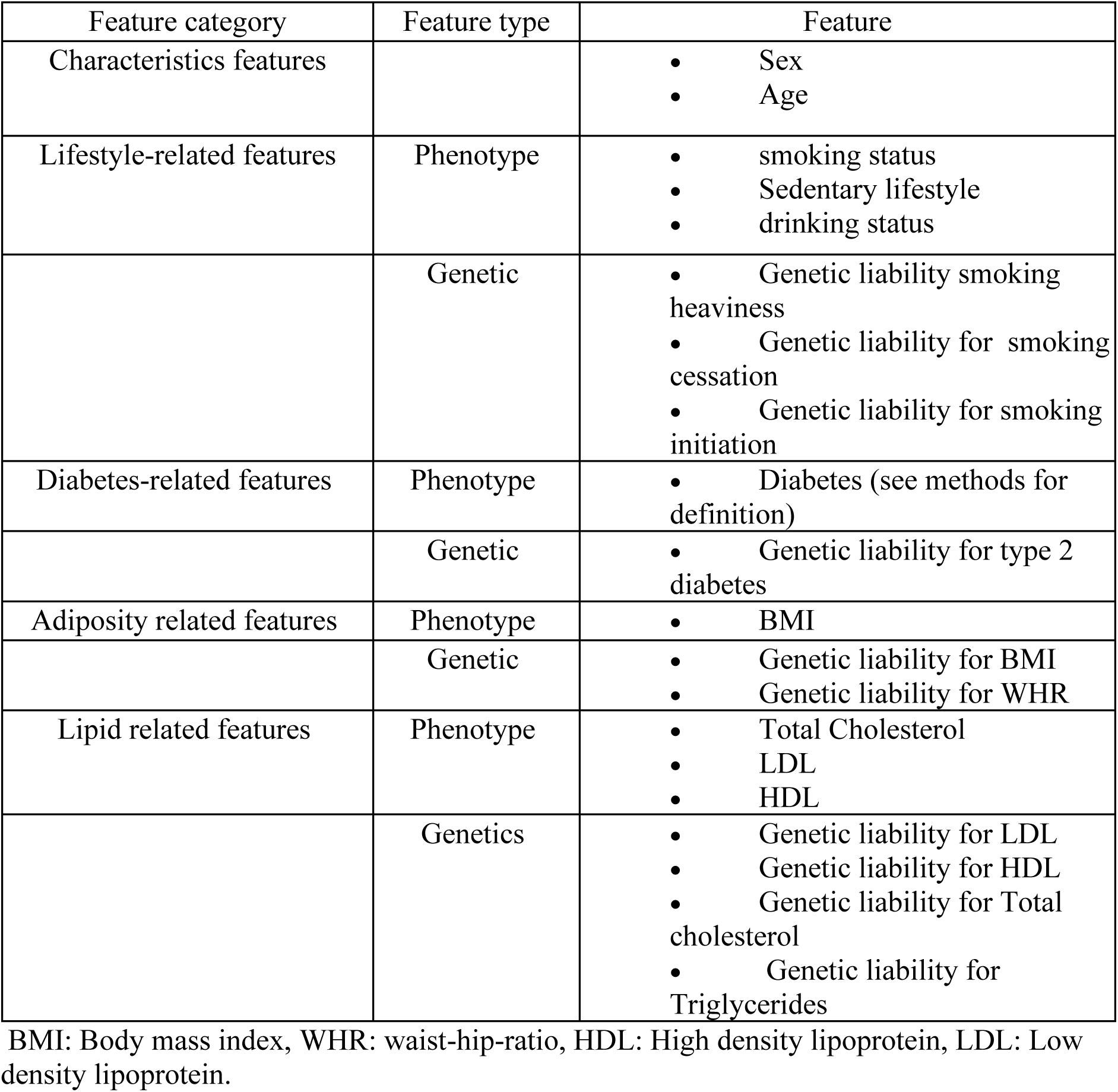
Features included in the machine learning algorithm.

**Table 3:**
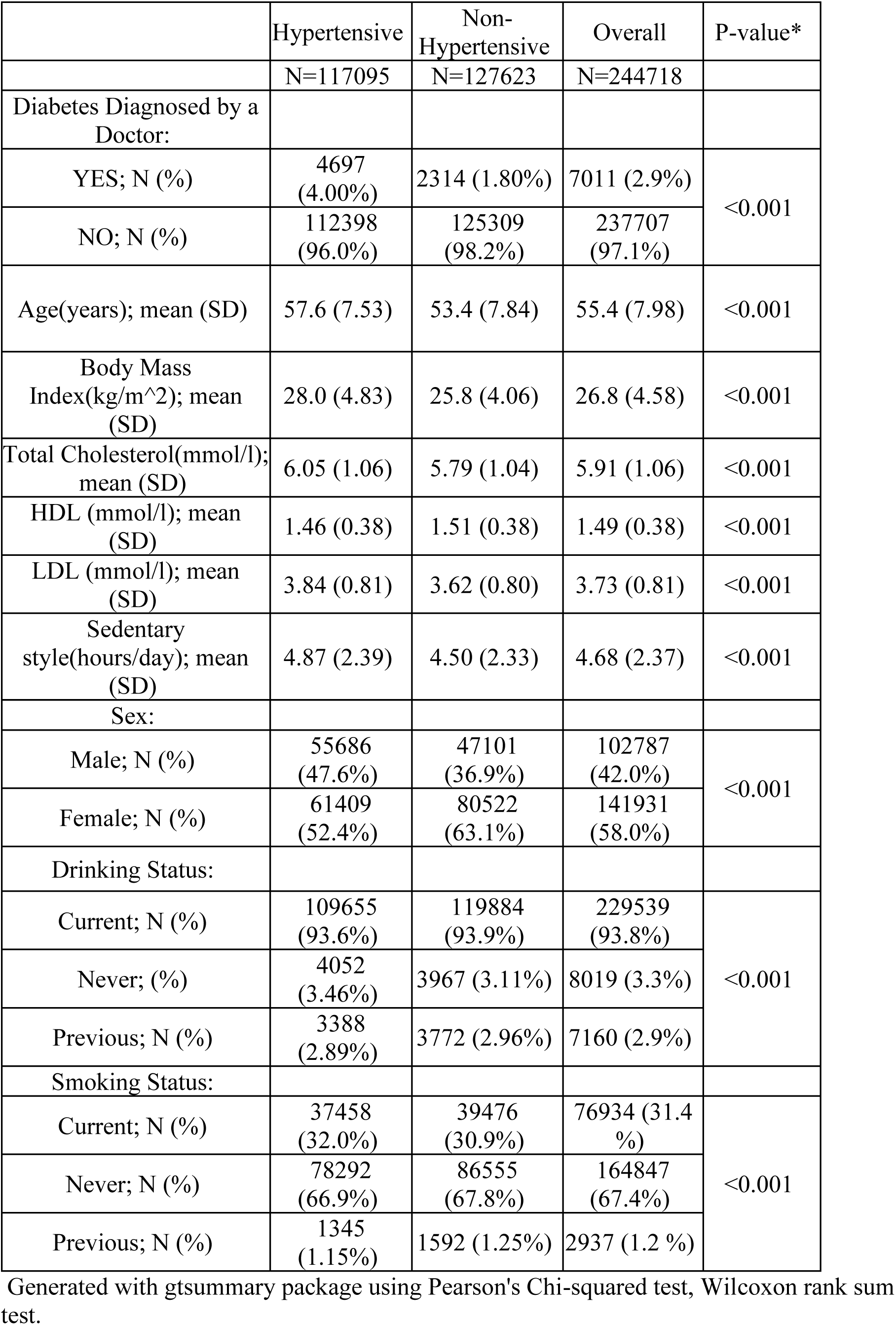
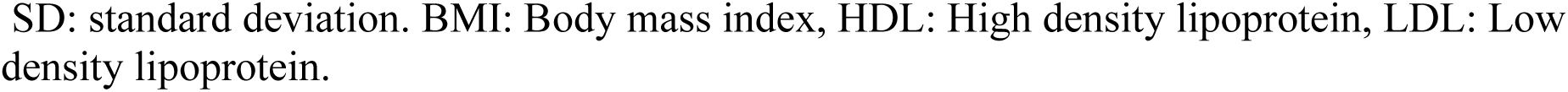
Baseline characteristic of the UK biobank participants within the overall sample and hypertensive subgroups.

|  | Hypertensive | Non-Hypertensive | Overall | P-value* |
| --- | --- | --- | --- | --- |
|  | N=117095 | N=127623 | N=244718 |  |
| Diabetes Diagnosed by a Doctor: |  |  |  |  |
| YES; N (%) | 4697 (4.00%) | 2314 (1.80%) | 7011 (2.9%) | <0.001 |
| NO; N (%) | 112398 (96.0%) | 125309 (98.2%) | 237707 (97.1%) |  |
| Age(years); mean (SD) | 57.6 (7.53) | 53.4 (7.84) | 55.4 (7.98) | <0.001 |
| Body Mass Index(kg/m<sup>2</sup>); mean (SD) | 28.0 (4.83) | 25.8 (4.06) | 26.8 (4.58) | <0.001 |
| Total Cholesterol(mmol/l); mean (SD) | 6.05 (1.06) | 5.79 (1.04) | 5.91 (1.06) | <0.001 |
| HDL (mmol/l); mean (SD) | 1.46 (0.38) | 1.51 (0.38) | 1.49 (0.38) | <0.001 |
| LDL (mmol/l); mean (SD) | 3.84 (0.81) | 3.62 (0.80) | 3.73 (0.81) | <0.001 |
| Sedentary style(hours/day); mean (SD) | 4.87 (2.39) | 4.50 (2.33) | 4.68 (2.37) | <0.001 |
| Sex: |  |  |  |  |
| Male; N (%) | 55686 (47.6%) | 47101 (36.9%) | 102787 (42.0%) | <0.001 |
| Female; N (%) | 61409 (52.4%) | 80522 (63.1%) | 141931 (58.0%) |  |
| Drinking Status: |  |  |  |  |
| Current; N (%) | 109655 (93.6%) | 119884 (93.9%) | 229539 (93.8%) | <0.001 |
| Never; (%) | 4052 (3.46%) | 3967 (3.11%) | 8019 (3.3%) |  |
| Previous; N (%) | 3388 (2.89%) | 3772 (2.96%) | 7160 (2.9%) |  |
| Smoking Status: |  |  |  |  |
| Current; N (%) | 37458 (32.0%) | 39476 (30.9%) | 76934 (31.4 %) | <0.001 |
| Never; N (%) | 78292 (66.9%) | 86555 (67.8%) | 164847 (67.4%) |  |
| Previous; N (%) | 1345 (1.15%) | 1592 (1.25%) | 2937 (1.2 %) |  |

Generated with gtsummary package using Pearson's Chi-squared test, Wilcoxon rank sum test.
SD: standard deviation. BMI: Body mass index, HDL: High density lipoprotein, LDL: Low density lipoprotein.

### Stage one models

In stage one models (Figure 2), the traditional features included the baseline characteristics as the predicting features (i.e. age, sex, BMI, diabetes, smoking status, drinking status, total cholesterol, HDL, LDL, and sedentary lifestyle; see methods). The saturated models included ten genetic liabilities in addition to the traditional features.

The traditional models (Table 4 and Figure 3 top panel) achieved AUCs of 0.70 (95% CI = 0.70, 0.71) using the Random Forest method (Table 4 and Figure 3 top left panel) and 0.72 (95%CI= 0.71, 0.72) using Neural Network (Table 4 and Figure 3 top right panel). The calibrations measured by Spiegelhalter’s z score were 1.03 (P-value= 0.30, calibration slope=0.98) for Random Forest (Table 4 and Supplementary Figure S2 Top left panel) and −14.39 (P-value= 6.4×10^−47^, calibration slope=1.18) for Neural Network (Table 4 and Supplementary Figure S2 top right panel). The addition of the genetic liabilities resulted in a slight improvement in the AUC only for Random Forest (AUC=0.71; Table 4 and Figure 3 bottom left panel).

**Figure 3.**
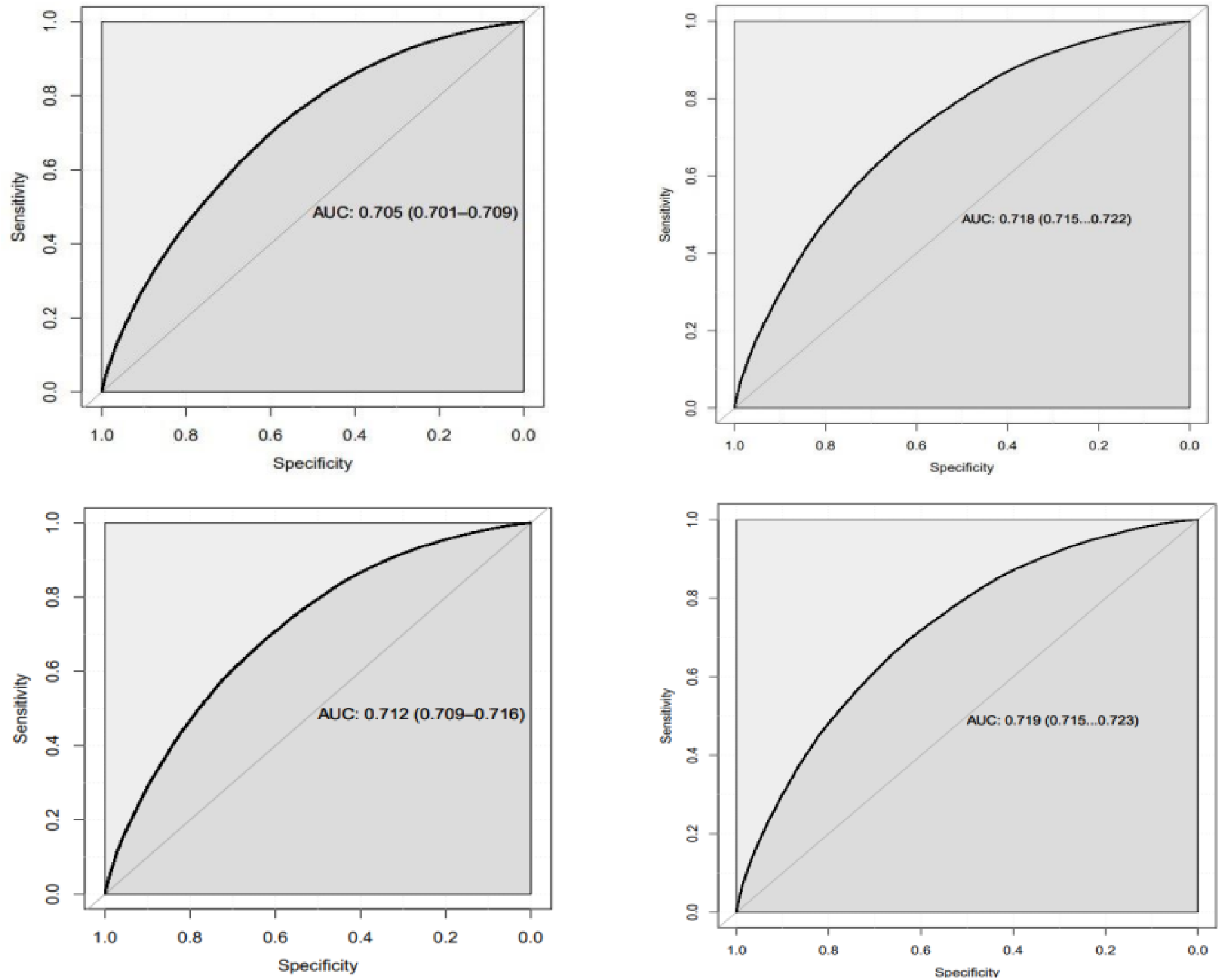
Top panel shows ROC plot for traditional Models. The figure shows the area under the curve (AUC) for both traditional Random Forest (top left panel) and traditional Neural Network (top right panel). **The bottom panel shows ROC for saturated Models.** The figure shows the AUC for saturated Random Forest (bottom left panel) and saturated Neural Network (bottom right panel).

**Table 4:** Discrimination and calibration results of the models applied to the testing set.

| Classification models | Numb of Features | R<sup>2</sup> | AUC% (95%CI) | Brier Score | Spiegelhalter z score | Spiegelhalter P-value | Slope | Intercept |
| --- | --- | --- | --- | --- | --- | --- | --- | --- |
| Models without Genetic Liabilities (Traditional models) |  |  |  |  |  |  |  |  |
| Random Forest | 10 | 0.17 | 0.70<br>(0.70,0.71) | 0.22 | 1.03 | 0.30** | 0.98 | 0.04 |
| Neural Network | 10 | 0.19 | 0.72<br>(0.71,0.7) | 0.21 | -14.39 | 6.4×10<sup>-47</sup> | 1.18 | 0.08 |
| Models with Genetic Liabilities (Saturated model= Traditional model + Genetic liabilities) |  |  |  |  |  |  |  |  |
| Random Forest | 20 | 0.18 | 0.71<br>(0.71,0.72) | 0.22 | -5.64 | 1.7×10<sup>-08</sup> | 1.06 | -0.04 |
| Neural Network | 20 | 0.19 | 0.72<br>(0.71,0.72) | 0.21 | -14.44 | 3.0×10<sup>-47</sup> | 1.18 | 0.07 |
| Random Forest as feature selection method |  |  |  |  |  |  |  |  |
| Random Forest | 10 | 0.17 | 0.71 0<br>(70,0.71) | 0.22 | 0.10 | 0.92** | 0.99 | -0.04 |
| Neural Network | 10 | 0.18 | 0.72<br>(0.71,0.72) | 0.21 | -15.51 | 3.1×10<sup>-54</sup> | 1.20 | -0.09 |
| Neural Network as feature selection method |  |  |  |  |  |  |  |  |
| Random Forest | 10 | 0.16 | 0.70<br>(0.70,0.71) | 0.22 | -0.44 | 0.66** | 1.00 | -0.04 |
| Neural Network | 10 | 0.17 | 0.71<br>(0.70,0.71) | 0.22 | -13.80 | 1.6×10<sup>-43</sup> | 1.18 | -0.08 |

Traditional model: age, sex, body mass index, diabetes, Smoking Status, Drinking Status, Total Cholesterol, HDL, LDL, and Sedentary Lifestyle. Discrimination is measured by the AUC. The Brier score is a combined measure of discrimination and calibration. Calibration is measured by the Spiegelhalter z test, logistic slope, and intercept. \*\*P-value >0.05(test is not significant) good calibration, AUC: Area under the curve, HDL: High density lipoprotein, LDL: Low density lipoprotein.

The saturated models showed poor calibration using both Random Forest and Neural Network methods (Table 4). The saturated Random Forest model showed a Spiegelhalter’s z score of −5.64 (P-value= 1.7×10^−08^, calibration slope=1.06; Table 4 and Supplementary Figure S2 bottom left panel). The saturated Neural Network model showed a Spiegelhalter’s z score of −14.44 (P-value= 3.0×10^−47^, calibration slope=1.18; Table 4 and Supplementary Figure S2 bottom Right panel)

### Stage two models

Feature selection (Figure 2) using Random Forest identified age as the most important classifying feature for hypertension, followed by sex, BMI, total cholesterol, LDL, sedentary lifestyle, HDL, total cholesterol genetic liability, LDL genetic liability, and smoking status. (Supplementary Figure S1 left panel). Feature selection using Neural Network identified HDL as the most important feature followed by total cholesterol, LDL, sedentary lifestyle, LDL genetic liability, BMI, total cholesterol genetic liability, age, WHR genetic liability, and HDL genetic liability (Supplementary Figure S1 right panel).

The first model (i.e. model built with the important features selected using Random Forest and classified using Random Forest; see methods) achieved an AUC of 0.71 (95 %Cl= 0.70, 0.71; Table 4 and Figure 4 Top left panel). The model was well-calibrated showing a Spiegelhalter’s z score of 0.10 (P-value= 0.92, calibration slope=0.99; Supplementary Figure S3 Top left panel).

**Figure 4.**
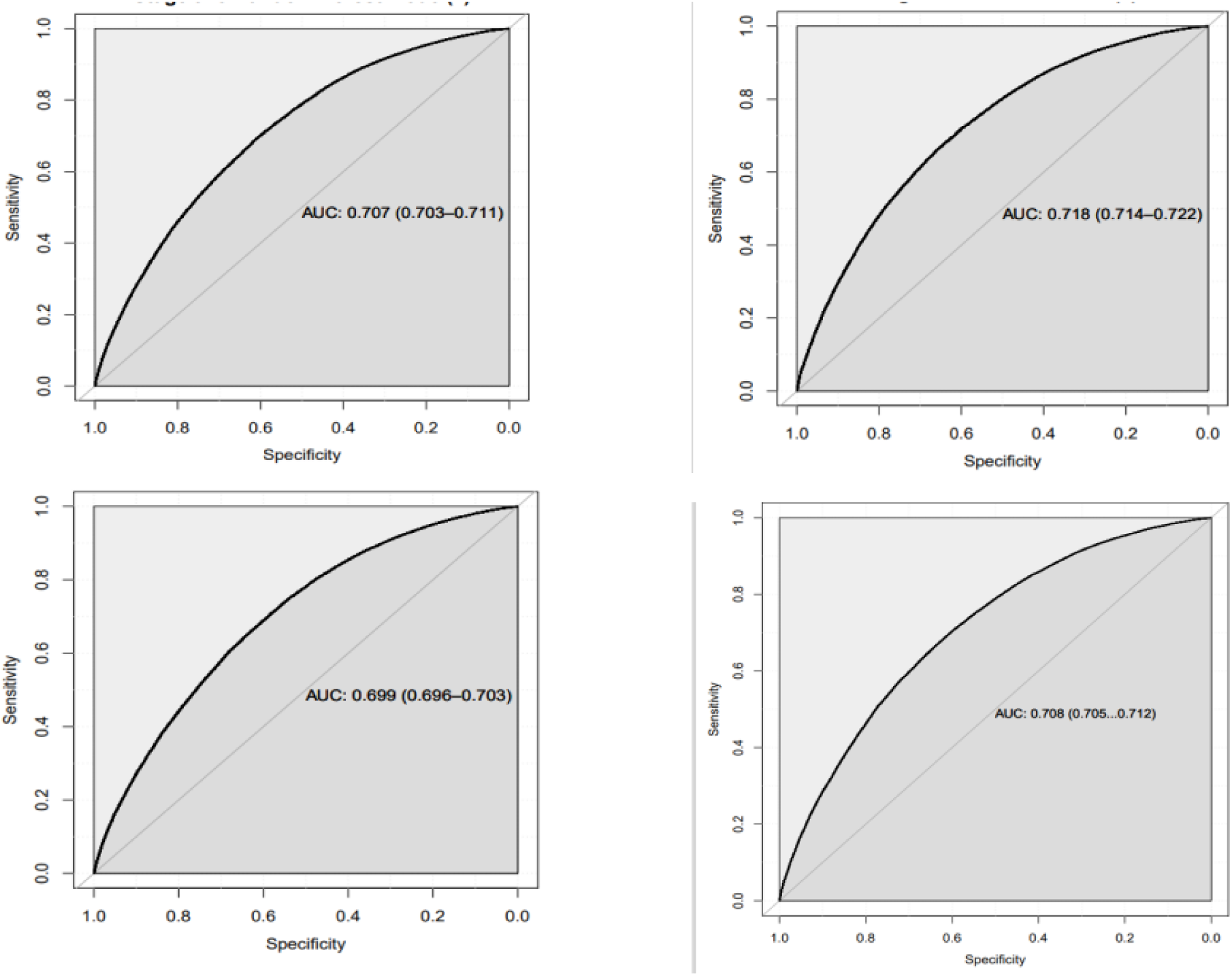
Top panel shows the ROC plot for models created with features selected by a saturated Random Forest. The figure shows the area under the curve (AUC) for Random Forest (top left panel) and Neural Network (top right panel). **The bottom panel shows ROC plot for models created with features selected by saturated Neural Network.** The figure shows the AUC for Random Forest (bottom left panel) and Neural Network (bottom right panel).

The second model (i.e. the model built with the important features selected using Random Forest and classified using Neural Network) achieved and AUC of 0.72 (95 %CI= 0.71, 0.72; Table 4 and Figure 4 Top right panel) but was poorly calibrated showing Spiegelhalter’s z score of −15.51 (P-value= 3.1×10^−54^, calibration slope=1.20; Supplementary Figure S3 Top right panel).

The third model (i.e. model built with the feature selection using Neural Network and classified using Random Forest method) achieved an AUC of 0.70 (95 %Cl= 0.70, 0.71; Table 4 and Figure 4 bottom left panel). The model was well-calibrated (Spiegelhalter’s z score −044, P-value= 0.66, calibration slope=1.00; Table 4 and Supplementary Figure S3 bottom left panel).

The fourth model (i.e. model with the feature selection and classified using Neural Network method) achieved AUC of 0.71 (95 %CI= 0.71, 0.72; Table 4 and Figure 4 bottom right panel) but was poorly calibrated (Spiegelhalter’s z score= −13.80, P-value= 1.6×10^−43^, calibration slope=1.18; Table 4 and Supplementary Figure S3 bottom right panel).

Three models with Random Forest classifier including one from stage one and two from stage two analysis were identified as well-calibrated (Figure 2). These models were included in the reclassification index analysis where the stage one model was used as the reference (i.e. model including all traditional features and using Random Forest as classifier; Figure 2). The model built with both feature selection and classification using Random Forest (Table 5) showed a slightly improved reclassification compared with the reference model indicated by a net reclassification index of 0.06 (95% CI= 0.05,0.08; Table 5). The model showed an integrated discrimination index of 1.7 ×10^−03^ (95% CI= 9.0×10^−04^, 2.5×10^−03^; Table 5). Conversely, the model built with the feature selection using Neural Network and classified using Random Forest method showed a deteriorated reclassification compared the reference model indicated by a net reclassification index of −010 (95% CI= 0.12,0.09; Table 5). The model showed an integrated discrimination index of −0.01(95% CI=-9.3×10^−04^, −0.01; Table 5).

**Table 5:** Net reclassification and integrated discrimination index.

| Feature selection Method | Classification Method | NRI<sup>>0</sup><br>(95 %CI)<br>P-value | IDI<br>(95 %CI)<br>P-value |
| --- | --- | --- | --- |
| None | *Random Forest | Ref | Ref |
| Random Forest | Random Forest | 0.06<br>(0.05,0.08)<br>P-value<0.00001 | $1.7 \times 10^{-03}$<br>($9.0 \times 10^{-04}$, $2.5 \times 10^{-03}$)<br>P-value=$1.0 \times 10^{-05}$ |
| Neural Network | Random Forest | -0.10<br>(-0.12, -0.09)<br>P-value<0.00001 | -0.01<br>(- $9.3 \times 10^{-04}$, -0.01)<br>P-value<0.00001 |

\*The traditional Random Forest model was chosen as a reference model. NRI<sup>>0</sup>: Continuous Net Reclassification Index, IDI: Integrated Discrimination Index, CI: Confidence Interval, Ref: Reference

## Discussion

This is the first and large-scale research that has been carried out on hypertension classification using machine learning that investigates the prediction value of a combination of genetic liabilities for type 2 diabetes, adiposity traits, lipids traits, and smoking traits in a single model. Aided by machine learning, we used a European dataset with 244,718 participants from the UK Biobank and identified the best integrated predictive models for classification of hypertension. We found that incorporating multiple genetic risk factors into prediction models could lead to minor but statistically significant improvement in the classification ability and reclassification of the models beyond conventional risk factors. Of all the genetic liabilities we considered, those estimated for total cholesterol and LDL cholesterol were identified to be a combination that could improve the classification of hypertension compared with the model without any genetic factors. This is the first study that identifies the predictive value of genetic liability of lipid traits in hypertension classification. Several cohort studies have found a link between high cholesterol levels (72, 73) as well as dyslipidemia (74, 75) and an increased risk of developing hypertension. Dyslipidemia is known to impair the functional and structural features of the arteries and cause atherosclerosis (76). These changes may compromise blood pressure control, predisposing individuals with dyslipidemia to hypertension.

Previous literature has only described traditional statistical techniques and machine learning models for hypertension mainly with non-genetic risk factors (18–23, 77). The studies that included genetic risk factors were limited to simply using single SNPs at a time (78, 79) and gene expression (80). In addition, the studies that utilised machine learning models (25) and included genetic liabilities were limited to one genetic liability at a time. Our study is unique in a sense that it incorporated multiple polygenic liabilities using machine learning to investigate a more integrated strategy in the classification of hypertension. Niu and colleagues incorporated a genetic liability component within machine learning models for hypertension (25) in which the authors used three machine learning models including Random Forest and Neural network to predict hypertension in rural China. The models included an Asian ancestry hypertension polygenic risk score (PRS) calculated using 13 single-nucleotide polymorphisms (SNPs). Our study included 10 various genetic liabilities incorporating a total of 883 SNPs. The study by Niu and colleagues found that including hypertension PRS in the models improved hypertension incidence prediction and risk reclassification (AUC_Random Forest_=0.84; AUC_Neural Network_=0.80). Our study took a different approach in terms of the type of genetic liabilities used. Instead of incorporating hypertension genetic liability, we included genetic liabilities for risk factors associated with hypertension and CVD (see methods). Compared with the study by Niu and colleagues, our study achieved a lower performance. Also, in terms of the net reclassification improvement in prediction value, our study showed only a marginal improvement whereas the study by Niu and colleagues showed an improvement of up to 4.7% in prediction value. This implies that incorporating genetic factors relating to the risk factors of hypertension may not be as promising as incorporating the hypertension genetic liability itself. However, it should be noted that our study investigated a large-scale European ancestry population and the study by Niu and colleagues investigated a population of Asian ancestry in rural China (The Henan Rural Cohort Study). These two populations have significant differences in their genetic make-up. Another reason for the observed differences could be environmental exposures and lifestyle variables, which can play a role in modifying the expression or impact of these genetic variants on phenotypes across populations (81, 82).

Our feature selection approach was successful in creating machine learning models that slightly improved the classification of hypertension. However, this came at the price of excluding clinically relevant features (e.g. diabetes mellitus and drinking status). We used a specific definition for diabetes (diabetes diagnosed by a doctor, or use of diabetic medication, or Hb1Ac ≥48 mmol/mol or glucose level ≥ 7.0 mmol/dl) (40). However, literature shows that diabetes mellitus and hypertension may co-exist, and it is not exactly clear which of the two precedes the other (83, 84). The observation that our machine learning feature selection approach did not prioritize diabetes as an important feature in classifying hypertension may align with the existing inquiries in the literature regarding the extent to which diabetes influences the development of hypertension, or conversely (85).

A strength of our study is in the novelty of the approaches used including (1) the use of machine learning to build a prediction model of hypertension in European setting, (2) in testing various methods of feature selection to identify the best performing set of predictive features and to ensure that the features included in the final model were robust and the model was well-calibrated, and (3) addition of multiple genetic liabilities in one single prediction model to identify the best performing classification model. In our integrated genetic approach, we included multiple genetic liabilities comprising a large number of SNPs within 10 genetic liabilities and allowed machine learning to identify the best pattern of feature combination in terms of model performance and accuracy. This gave us a comprehensive picture of the effectiveness of various genetic liabilities in comparison with each other and hypertension risk factors. Another strength is in the use of the large sample size of the UKB that allowed us to develop a large training set comprising 171,304 participants. This is beneficial in detecting the true effect of risk factors on outcomes, reducing bias, and making risk predictions in the testing set more reliable (86, 87). Our study contributes to the ongoing research on the potential role of genetic liabilities in risk prediction of complex diseases (88–90).

A limitation of our research is that the UK biobank data is imbalanced in terms of the ratio of cases and controls and as a result, our sample included 10,528 more controls than the cases. The training set included 7,370 more controls than cases. Models trained on imbalanced datasets may become biased towards the dominant class, predicting the minority class incorrectly (48). To address the imbalance in the dataset and minimise error, we utilised an over-sampling approach to balance the sample (91). This may introduce noise into the synthetic sample in the dataset, resulting in some level of bias remaining in the models (92). In this study, we also used an integrative approach and included multiple features in a machine learning model with the 5-fold cross-validation techniques which enabled us to evaluate the performance of the model on multiple subsets of the training set. However, some residual overfitting caused by the possible complexity of the model might still exist in the data despite the application of the cross-validation technique (93). To overcome this issue caused by potentially irrelevant features, we performed a sensitivity analysis and used a feature selection technique to focus on the most important features. Another limitation of our study is that the population investigated was of European ancestry, which limits the generalizability of our findings to other ethnicities or ancestries. The non-European ancestral groups within the UKB have very small sample size which limits their statistical power to be used for genetic studies and specifically using machine learning methods that need partitioning data into training and testing sets. We propose that incorporating a more diverse participant pool and conducting studies across diverse populations in future work could potentially improve the generalizability and robustness of predicting hypertension using genetic liabilities.

## Conclusions

Our research highlighted that, out of the 10 genetic liabilities considered in our study, genetic liability for two lipids (total cholesterol and LDL) was found to add value to the classification of hypertension within a European ancestry population. The inclusion of these two genetic liabilities in the Random Forest model slightly improved the hypertension risk discrimination as well as risk reclassification for individual participants beyond the conventional factors. Incorporating multiple genetic liabilities in machine learning-based models is proposed for future studies which might identify complex patterns within the data.

## Author Contributions

Conceptualization, R.P.; Data curation, Formal analysis, G.M.; Funding acquisition, R.P.; Investigation, G.M.; Methodology, G.M., R.P.; Project administration, R.P.; Resources, R.P.; Supervision, R.P; Writing— original draft, G.M.; Writing—review & editing, G.M., and R.P. All authors have read and agreed to the published version of the manuscript.

## Funding

R.P. holds a fellowship supported by the Rutherford Fund from the Medical Research Council (MR/R026505/1 and MR/R026505/2). UKB genotyping was supported by the British Heart Foundation (grant SP/13/2/30111) for Large-scale comprehensive genotyping of UKB for cardiometabolic traits and diseases: UK CardioMetabolic Consortium.

## Institutional Review Board Statement

The study was conducted in accordance with the Declaration of Helsinki and approved by the Institutional Review Board (or Ethics Committee) of Brunel University London, College of Health, Medicine, and Life Sciences (27684-LR-Jan/2021-29901-1).

## Informed Consent Statement

Informed consent was obtained from all subjects involved in the study.

## Data Availability Statement

Not applicable.

## Data Availability

All data produced in the present study are available upon reasonable request to the authors

## Acknowledgments

This study has been performed using the UK Biobank application 60549.

## Conflicts of Interest

The authors declare no conflict of interest.

## Supplementary Materials

Supplementary Excel Table1-Supplementary Excel Table10: List of genetic variants summary statistics used to construct the genetic risk scores.

Supplementary Table 1–Supplementary Table 4

Supplementary Figure 1–Supplementary Figure 3

## Notes

### Competing Interest Statement

The authors have declared no competing interest.

### Funding Statement

This study did not receive any funding

### Author Declarations

Division of Biomedical Sciences, Department of Life Sciences, College of Health and Life Sciences, Brunel University London

