## Supplementary material for "Using machine learning to evaluate the value of genetic liabilities in classification of hypertension within the UK Biobank"

**How Genetic Liability Predicts the Occurrence of Hypertension Using Machine Learning Methods**

**Supplementary Information**

**Supplementary Table 1:** Overview of the association analysis between study predictors and hypertension based on univariable logistic regression analysis.

| Characteristic | N | OR^1^ | 95% CI^1^ | p-value |
| --- | --- | --- | --- | --- |
| Diabetes Mellitus (yes) | 244,718 | 2.26 | 2.15, 2.38 | <0.001 |
| Sex (Male) | 244,718 | 1.55 | 1.53, 1.58 | <0.001 |
| Age | 244,718 | 1.07 | 1.07, 1.07 | <0.001 |
| BMI | 244,718 | 1.12 | 1.12, 1.12 | <0.001 |
| Smoking Status | 244,718 |  |  |  |
| Current | reference | 1.0 | — |  |
| Never |  | 1.15 | 1.12, 1.18 | <0.001 |
| Previous |  | 1.32 | 1.28, 1.36 | <0.001 |
| Drinking Status | 244,718 |  |  |  |
| Current | reference | 1.0 | — |  |
| Never |  | 1.12 | 1.07, 1.17 | <0.001 |
| Previous |  | 0.98 | 0.94, 1.03 | 0.4 |
| Total Cholesterol | 244,718 | 1.27 | 1.26, 1.28 | <0.001 |
| HDL | 244,718 | 0.72 | 0.70, 0.73 | <0.001 |
| LDL | 244,718 | 1.39 | 1.37, 1.40 | <0.001 |
| Sedentary Lifestyle | 244,718 | 1.07 | 1.07, 1.07 | <0.001 |
| ^1^OR = Odds Ratio, CI = Confidence Interval | | | | |

**Supplementary Table 2:** Overview of the association analysis between study predictors and hypertension based on multivariable logistic regression analysis.

| Characteristic | N | OR^1^ | 95% CI^1^ | p-value |
| --- | --- | --- | --- | --- |
| Diabetes Mellitus(yes) | 244,718 | 1.56 | 1.48, 1.65 | <0.001 |
| Sex(male) | 244,718 | 1.70 | 1.67, 1.73 | <0.001 |
| Age | 244,718 | 1.07 | 1.07, 1.08 | <0.001 |
| BMI | 244,718 | 1.12 | 1.12, 1.13 | <0.001 |
| Smoking Status | 244,718 |  |  |  |
| Current | reference | 1.0 | — |  |
| Never |  | 1.11 | 1.08, 1.14 | <0.001 |
| Previous |  | 1.07 | 1.04, 1.10 | <0.001 |
| Drinking Status | 244,718 |  |  |  |
| Current | reference | 1.0 | — |  |
| Never |  | 0.98 | 0.94, 1.03 | 0.5 |
| Previous |  | 0.93 | 0.88, 0.97 | 0.003 |
| Total Cholesterol | 244,718 | 1.66 | 1.59, 1.73 | <0.001 |
| HDL | 244,718 | 0.91 | 0.87, 0.94 | <0.001 |
| LDL | 244,718 | 0.63 | 0.60, 0.66 | <0.001 |
| Sedentary Lifestyle | 244,718 | 1.01 | 1.01, 1.01 | <0.001 |
| ^1^OR = Odds Ratio, CI = Confidence Interval | | | | |

**Supplementary Table 3:** Optimal metrics identified for in the Random Forest models for the classification of hypertension based on the training data.

| Model | Number of Features | ML technique/ set | Outcome | Mtry | OOB Prediction error (Brier s.) |
| --- | --- | --- | --- | --- | --- |
| Traditional Random Forest | 10 | Random Forest/training set | hypertension | 3 | 0.22 |
| Saturated Random Forest | 20 | Random Forest/training set | hypertension | 4 | 0.22 |
| Stage 2 Random Forest ^†^ | 10 | Random Forest/training set | hypertension | 3 | 0.22 |
| Stage 2 Random Forest ^† †^ | 10 | Random Forest/training set | hypertension | 3 | 0.22 |

The machine learning technique for this table is Neural Network and the results are based on the training set. Results are from the ranger function within the ranger R package.

Traditional model: Models without genetic Liabilities (Baseline Models).

Saturated model: Models including genetic liabilities.

†: Model built with top ten important features selected by Saturated Random Forest.

† †:  Model built with top ten important features selected by Saturated Neural Network.

Mtry: the number of variables to randomly sample as candidates at each split.

ML: machine learning.

OOB: out of bag.

**Supplementary Table 4:** Optimal metrics identified in the Neural Network models for the classification of hypertension based on the training data.

| Model | Number of Features | Number of Hidden layers | ROC (SD) | Sensitivity (SD) | Specificity (SD) |
| --- | --- | --- | --- | --- | --- |
| Traditional Neural Network | 10 | 5 | 0.70 (0.004) | 0.62 (0.006) | 0.67 (0.004) |
| Saturated Neural Network | 20 | 5 | 0.70 (0.004) | 0.62 (0.006) | 0.67 (0.006) |
| Stage 2 Neural Network ‡ | 10 | 5 | 0.70(0.004) | 0.62(0.007) | 0.67(0.006) |
| Stage 2 Neural Network ^‡‡^ | 10 | 5 | 0.69(0.004) | 0.61(0.007) | 0.66(0.007) |

The machine learning technique for this table is Neural Network and the results are based on the training set. Results are from the nnet function within the caret R package.

‡: Model built with top ten important features selected by Saturated Random Forest.

‡‡:  Model built with top ten important features selected by Saturated Neural Network.

ROC: receiver operating characteristic curve.

SD: Standard deviation.

**Supplementary Figure S1**
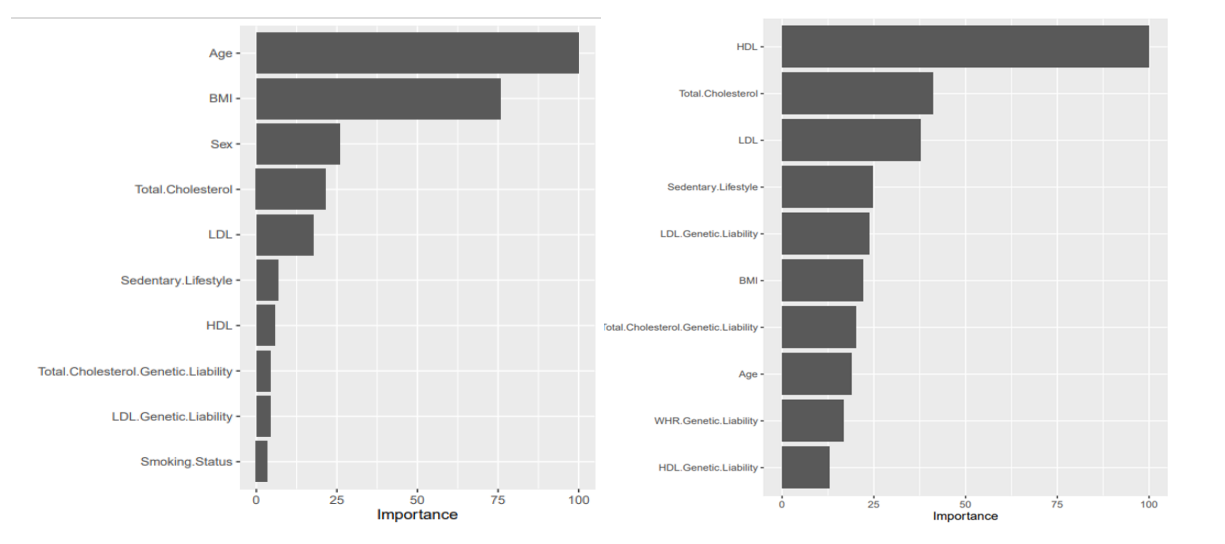

**Supplementary Figure S1: Top ten important Features selected by Random Forest (left panel) and by Neural Network (right panel).**

**Supplementary Figure S2**

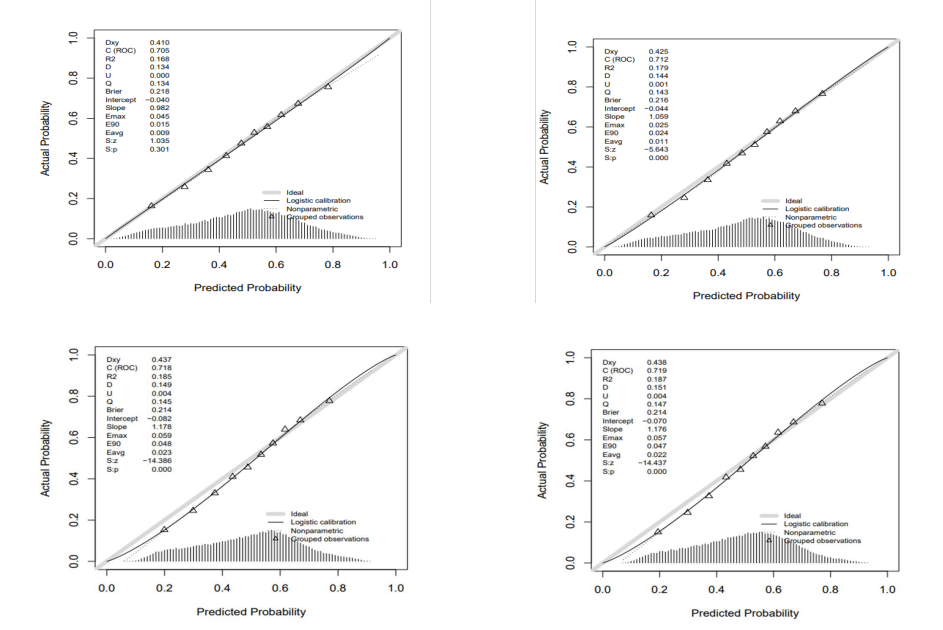

**Supplementary Figure S2: Calibration curve of stage one models**. Traditional Random Forest (top left panel) is miscalibrated due to overfitting and Saturated Random Forest (top right panel) is well calibrated. Traditional Neural Network (bottom left panel) is miscalibrated due to overfitting and Saturated Neural Network (bottom right panel) is miscalibrated due to overfitting. The solid grey line is ideal calibration, the solid black line is logistic calibration, the dotted line is a non-parametric calibration, and the triangular points are the grouped observations presence. The distribution plot of predicted probability is also displayed.

**Supplementary Figure S3:**

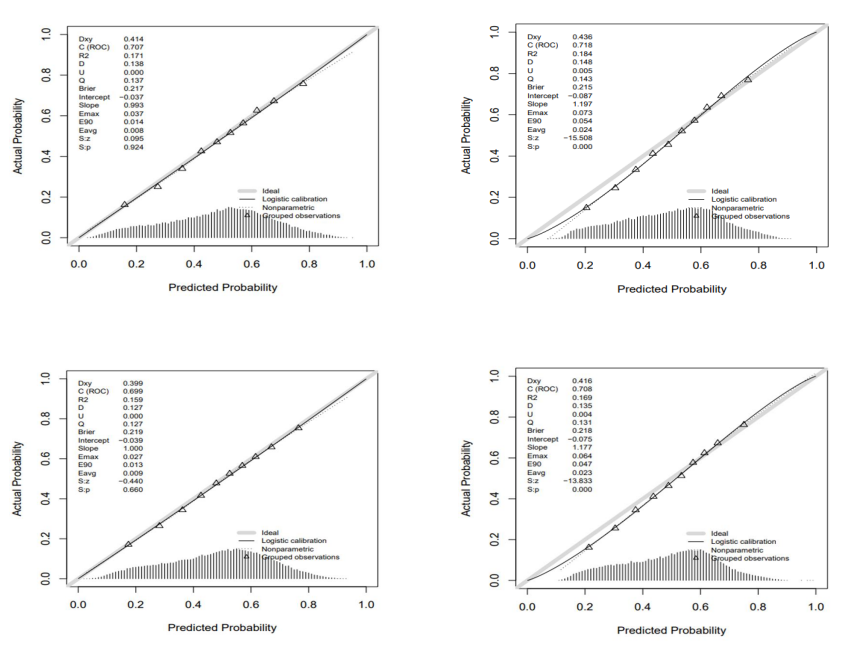

**Supplementary Figure S3: Calibration curve of stage two models.** Calibration curve of models created with features selected Random Forest (top panel). Random Forest (top left panel) is well calibrated and Neural Network (top right panel) is miscalibrated due to overfitting. Calibration curve of models created with features selected Neural Network (bottom panel). Random Forest (bottom left panel) is well calibrated and Neural Network (bottom right panel) is miscalibrated due to overfitting. The solid grey line is ideal calibration, the solid black line is logistic calibration, the dotted line is a non-parametric calibration, and the triangular points are the grouped observations presence. The distribution plot of predicted probability is also displayed.

**Supplementary Figure S4:**

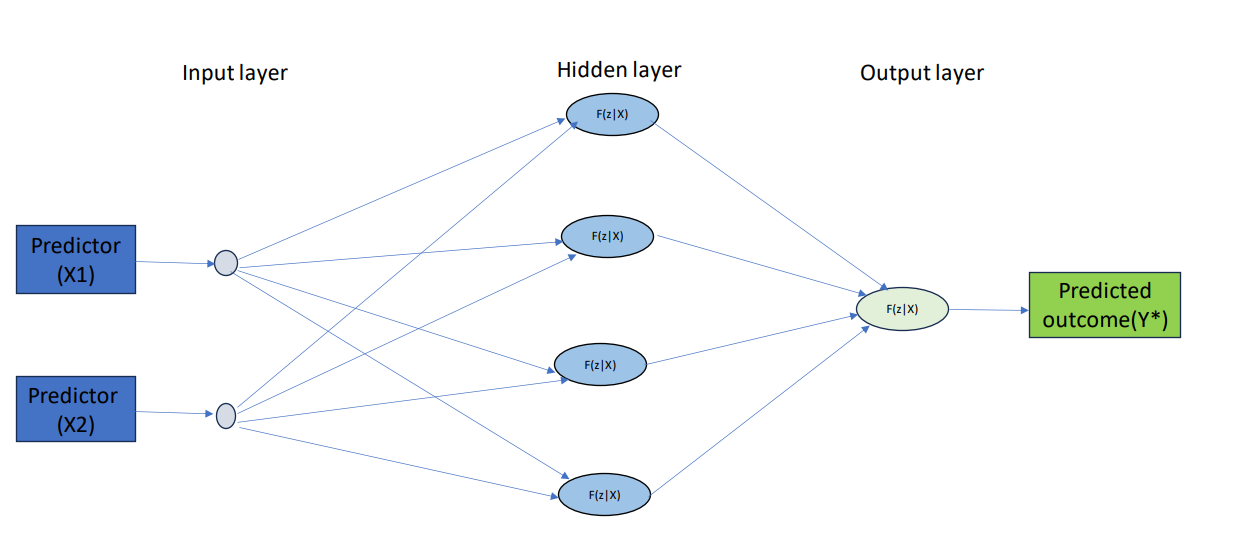

**Supplementary Figure S4: A simple architecture of Neural Network.**

F(z|x): Some black box function
